# Individuals with recurrent low back pain exhibit further altered trunk control in remission than when in pain

**DOI:** 10.1101/2020.10.15.20213488

**Authors:** Hai-Jung Steffi Shih, Linda Van Dillen, Jason Kutch, Kornelia Kulig

**Affiliations:** Division of Biokinesiology and Physical Therapy, University of Southern California, Los Angeles, CA, USA; Program in Physical Therapy, Washington University School of Medicine in St. Louis, St. Louis, MO, USA

**Keywords:** Gait, Muscle activity, Trunk Coordination, Vector Coding, Low Back Pain

## Abstract

**Background:** Movement alterations due to low back pain (LBP) could lead to long-term adverse consequences if they do not resolve after symptom subsides. This study aims to determine if altered trunk control associated with recurrent low back pain persists beyond symptom duration.

**Methods:** Twenty young adults with recurrent low back pain were tested once during an LBP episode and once in symptom remission, and twenty matched back-healthy participants served as controls. Participants walked on a treadmill with five prescribed step widths. Motion capture and surface electromyography were used to record frontal plane trunk kinematics and muscle activation. Thorax-pelvis coordination was calculated using vector coding technique, and bilateral longissimus activation and co-activation were analyzed.

**Findings:** Young adults with recurrent LBP exhibited a “looser” trunk control strategy in the frontal plane during gait that was persistent regardless of pain status across multiple step widths compared to back-healthy controls. This was demonstrated by a greater pelvis-only, less thorax-only coordination pattern, and decreased bilateral longissimus co-activation in individuals with recurrent LBP than controls. The looser trunk control strategy was further amplified when individuals with recurrent LBP were in symptom remission and exhibited greater trunk excursion and reduced in-phase coordination.

**Interpretation:** The amplification of aberrant movement during symptom remission may suggest that movement patterns or anatomical factors existing prior to the tested painful episode underlie the altered trunk control in individuals with recurrent LBP. The symptom remission period of recurrent LBP patients may be a critical window into clinical evaluation and treatment.

## Introduction

Low back pain (LBP) is highly prevalent, leads to global burden^1,2^, and often follows a recurrent course.^3^ About 80% of the population is affected by LBP, and up to 56% of the people that experience LBP exhibit a recurrence of symptoms within a year.^4,5^ LBP symptoms impact an individual’s quality of life, increase the risk of depression and risk of developing opioid dependence.^6–8^ The majority of LBP is non-specific, with no identifiable cause of symptoms which makes developing primary prevention programs challenging.^9^ Secondary prevention could greatly reduce personal and societal cost associated with LBP disability. The lack of established factors associated with future recurrence is a critical barrier to the development of effective intervention programs.^5^

Movement alterations attributed to a history of pain could lead to long-term adverse consequences if they persist beyond symptom duration. Studies have reported that a history of previous back pain episodes predicts future recurrences and persistence of symptoms, while the underlying mechanisms remain elusive.^5,10^ A plausible mechanism is that the motor system adapts to back pain and utilizes suboptimal control of movement which is beneficial in the short-term, but detrimental in the long-term.^11–16^ Recent literature reviews suggested that although many studies report a difference between persons with LBP and the controls, the direction may be dependent on the type of task and population.^17,18^ Both ends of the spectrum could have negative impact on spine health: a “tighter” trunk control (increased muscle activation, increased trunk stiffness and coupling, and decreased segmental movement) may lead to more compression loads causing joint and disc degeneration; a “looser” trunk control (decreased muscle activation, decreased trunk stiffness and coupling, and increased segmental movement) may lead to increased shear force resulting in soft tissue strains and joint irritation.^17,18^

There is evidence that altered motor control is not only present during painful periods, but also persists beyond symptom duration in persons with recurrent LBP.^19–23^ Asymptomatic participants with a history of recurrent LBP exhibit impaired dynamic balance^23^, increased trunk stiffness during a rapid load-release perturbation^19^, and a more in-phase thorax-pelvis coordination during running^20,24^ and walking^20^ compared to back-healthy controls. However, results regarding trunk coordination during walking are inconsistent, including more in-phase (tighter)^20^, more trunk-phase and less in-phase (looser)^21^, or no difference^24^ compared to controls in asymptomatic individuals with a history of LBP. Moreover, these studies tested individuals during symptom remission but did not provide a comparison with their behavior in pain. Therefore, current study aimed to test trunk control of a cohort of people with recurrent LBP in and out of an episode of LBP.

Pain is a potent stimulus and the body sees it as a great cost, therefore we learn to adapt to pain quickly, but may fail to further refine our strategies.^11^ Individuals with experimental and clinical LBP exhibit low movement variability once they have adopted a specific strategy to move with pain.^11,25^ In one experiment, lower muscle onset timing variability after adaptation to experimental pain was predictive of the failure to return to previous onset timing when pain subsided.^26^ These studies showed decreased movement variability for a given task; there is also evidence demonstrating the same altered movement strategy across different task demands.^27^ Patients with LBP may be less able to adapt to varying task demands due to a stereotypical movement strategy. Walking is a highly ecological task and requires active balance control in the frontal plane.^28^ Therefore, we designed an experiment where we systematically change step widths as a means to examine if individuals with LBP display an adaptable movement strategy.

The purpose of this study was to compare trunk control in and out of a painful episode in individuals with recurrent LBP, and to determine if the trunk control was altered when compared to back-healthy individuals. We achieved this by examining frontal plane trunk kinematics and longissimus muscle activation in response to different step widths during gait. Considering the predominant current literature, we hypothesized that individuals with recurrent LBP would display a tighter trunk control compared to controls regardless of whether or not they were in pain. We also expected a less adaptable movement strategy in response to different step widths in individuals with recurrent LBP compared to controls.

## Methods

### Participants

Sample size calculation based on the smallest partial η2 among all the variables of interest in our pilot data revealed that 14 participants in each group would be sufficient to reach 80% statistical power to detect differences between pain status within the LBP group, and between the LBP and control groups. Twenty participants with recurrent LBP and twenty matched back-healthy individuals completed the study (Supplementary figure 1).

Participants with recurrent LBP were included if they were between 18 and 45 years old, had pain that was localized to the area between the lower posterior margin of the rib cage and the horizontal gluteal fold for more than 6 months, but had less than half of the days in pain to differentiate from chronic representation.^29^ They also had to have episodes that limit function based on any item other than pain scoring ≥ 1 on the Oswestry Disability Index (ODI).^30^ They were excluded if they had a history of leg pain below the knee accompanying their LBP, pain that lasted more than 6 months in other body regions, a history of spine or lower extremity surgery, a radiological or clinical diagnosis of any spinal structural pathology, malignancy, infection, stenosis, or radiculopathy (based on neurological screening for sensory, motor, and reflex integrity), a history of diabetes mellitus that affected peripheral sensation, rheumatic joint disease, polyneuropathy, ankylosing spondylitis, active cancer, or current pregnancy. They were also excluded if they had any condition that would prevent completion of the experimental tasks or is known to severely affect balance or locomotion.

Back-healthy controls were age (± 5 yrs), sex, body mass index (BMI) (in the same category), and activity (± 15% metabolic equivalents per week based on the International Physical Activity Questionnaire) matched to the recurrent LBP group. They were included if they had no LBP that affected activity in the previous year, determined by self-report. They were excluded based on the same criteria as the recurrent LBP group.

Participants with recurrent LBP were tested twice, first when their pain had persisted for more than 24 hours at the level of ≥2/10 on a written numeric pain rating scale,^31^ and then when their pain was <1/10 for more than 24 hours. Given the high test-retest reliability (see results), the back-healthy group was only tested once. Participants gave written informed consent that was approved by the institutional review board of the University of Southern California.

### Instrumentation

Participants were instrumented with reflective markers placed on the lower extremity and trunk.^32^ Kinematic data were recorded by an 11-camera Qualysis motion capture system (Qualisys Inc., Gothenburg, Sweden) at 125 Hz. Surface electromyography (EMG) was collected using bipolar silver chloride electrodes with an interelectrode distance of 22 mm. Electrodes were placed on bilateral longissimus (2 fingers width lateral to the L3 spinous process surface).^33^ Data were collected using a wireless EMG system (Noraxon U.S.A, Inc., Scottsdale, AZ, USA) at 1500 Hz. A portable treadmill (PRO-FORM 505 CST, ICON Health & Fitness, Inc., Logan, UT, USA) was used for the walking trials.

### Experimental procedures

Participants were electronically screened for eligibility before testing in the laboratory. They underwent a clinical screening of scoliosis using the Adam’s forward flexion test, and a clinical screening of sensory, motor, and reflex function. They also completed the International Physical Activity Questionnaire^34,35^, and a medical history form that incorporated the minimal dataset recommended by the National Institutes for Health task force on research standards for chronic LBP.^29^ Individuals with recurrent LBP completed the ODI^30^, the Fear Avoidance Beliefs Questionnaire (FABQ)^36^, and a body pain diagram to indicate pain location^37^. They were also required to complete several visual analog scales (0-100 mm) for pain at rest and during walking. Leg length from the greater trochanter to the ground of the right leg was measured.

Participants then were given 3 minutes to familiarize themselves to the treadmill, after which a 30 second treadmill walking trial at 1.25 m/s was collected to determine preferred step width (PSW). Participants were then introduced to step widths with real-time visual feedback projected on the wall in front of the treadmill (Fig 1). Step width was calculated using marker data that was streamed real-time into a custom program in MATLAB (MathWorks, Inc., Natick, MA, USA). The averaged heel and 2^nd^ toe marker position was defined as the foot position, then step width was determined as the medial-lateral distance between foot positions at consecutive foot flat. Five different target step widths were prescribed, including 0.33, 0.67, 1, 1.33, and 1.67 × PSW. Participants were given a practice trial for each prescribed width, before completing a 30-second trial for each step width presented in a randomized order.

**Figure 1.**
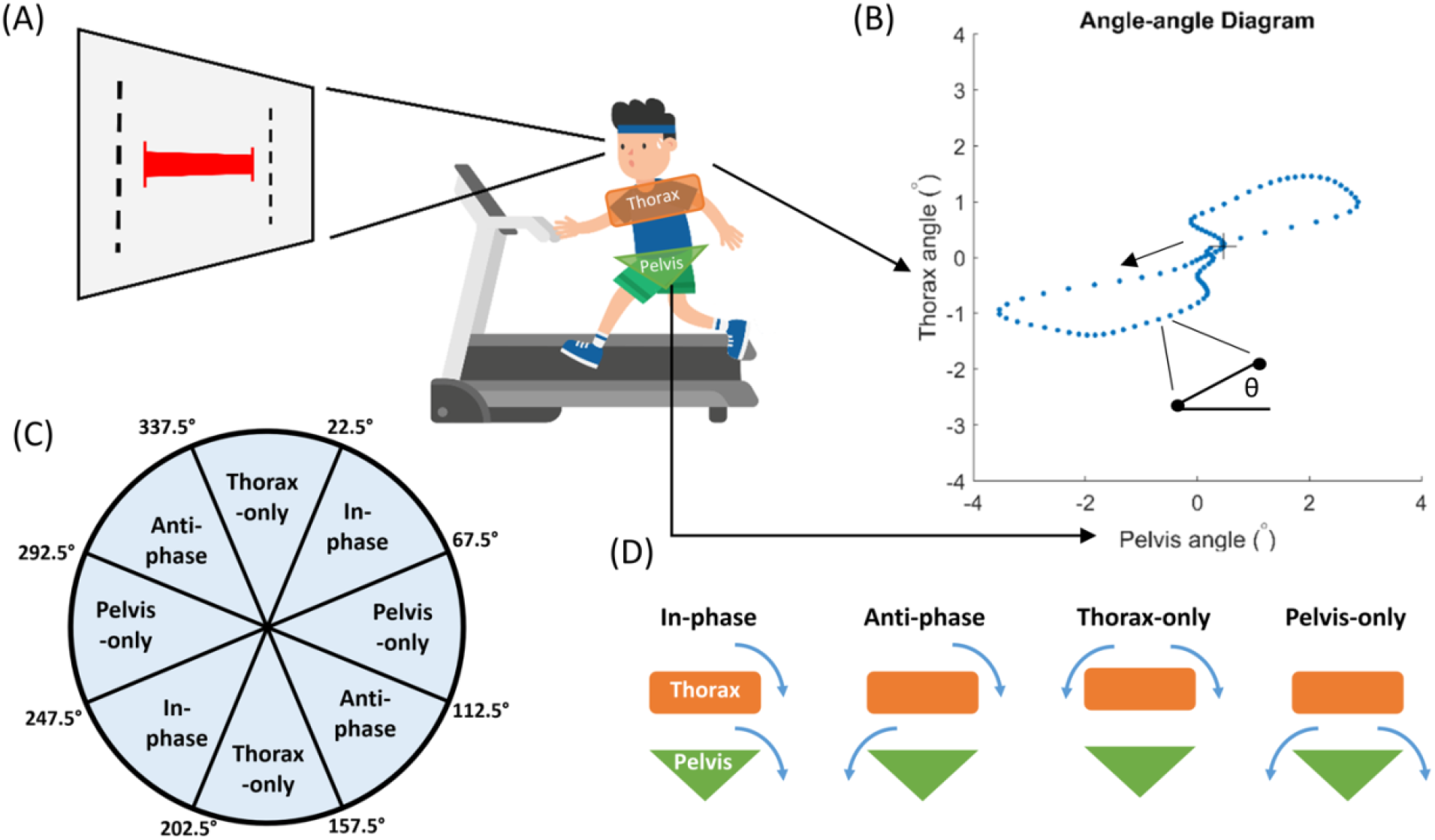
Experimental setup of treadmill walking with prescribed step widths. A visual feedback was projected on a wall in front of the treadmill, with a red horizontal bar representing participant’s actual step width and black vertical lines indicating the target width. Participants were instructed to match the width of a red bar to the black dotted lines. The thorax and the pelvis segments are illustrated. (B) A pelvis-thorax angle-angle diagram of a gait cycle in one representative participant. The “+” denotes right heel strike and the arrow indicate progression of movement. Coupling angle was defined as the vector angle of two consecutive points in time relative to the right horizontal. (C) Cutoffs for binning of the coupling angles into four coordination patterns. (D) Illustrations of the physical implication of the four coordination patterns. In-phase indicate that both segments are rotating to the same direction at similar rate; anti-phase indicate that the segments are rotating to the opposite direction at similar rate; thorax-only and pelvis-only indicate that the thorax or pelvis segment is rotating significantly faster than the other segment, or the other segment is hardly rotating.

### Data Analyses

Kinematic data were low-pass filtered at 10 Hz with a dual-pass 4^th^ order Butterworth filter. Gait events were identified based on the toe and heel marker position relative to the pelvis coordinate. Each gait cycle was time-normalized to 101 data points from right heel strike to the subsequent right heel strike. Each 30-second trial consisted of 25-35 strides, but only the last 20 strides were included in the analyses to represent steady state walking with the prescribed step widths.

The frontal plane trunk excursion and thorax-pelvis kinematic coordination were analyzed as this was the plane step width changes occur in. Trunk excursion was defined as the range of thorax relative to the pelvis rotation during gait. Thorax-pelvis kinematic coordination was calculated using vector coding.^38^ Specifically, an angle-angle diagram was first constructed with the pelvic angle on the horizontal axis and the thorax angle on the vertical axis, then the coupling angle was calculated as the angle of the vector between two adjacent data points in time relative to the right horizontal (Fig 1A). Coordination patterns were categorized as in-phase, anti-phase, thorax-only, and pelvic-only defined by coupling angles falling within each range indicated in Fig 1B (Fig 1B&C).

EMG data were bandpass filtered between 30 Hz and 500 Hz to avoid heart rate contamination with a dual-pass 4^th^ order Butterworth filter.^39^ The signal was then full-wave rectified and smoothed using a 100 ms moving window, and normalized to the averaged peak gait activation of a normal treadmill walking trial visual feedback. The peak longissimus EMG during the contralateral stance phase, where the greater activation occurred, was calculated. Bilateral co-activation for longissimus was determined as the average ratio between right and left muscle activation, where the less activated side was always the numerator.

### Statistical Analysis

Data were plotted for visualization and examined for normality, homoscedasticity, and outliers. Descriptive statistics were performed, and paired t-tests were used to compare participant characteristics, except for sex as the two groups were identical. The analyses were conducted in steps. First, all dependent variables were compared within the recurrent LBP group when they were in active pain (rLBP-A) versus in remission (rLBP-R). The dependent variables examined included the following: trunk excursion, 4 patterns of trunk coordination, peak longissimus activation, and bilateral longissimus co-activation. Data were analyzed using general mixed effect models. The independent variables were step width (here we used the actual step widths participants achieved, a continuous variable) and pain status (rLBP-A versus rLBP-R) as fixed effects, and subjects as a random effect. Second, if there were no significant effects of pain status or an interaction, then data for rLBP-A and rLBP-P were pooled (rLBP) and compared to the back-healthy control group (CTRL) using general mixed effect models with the same structure, but replacing pain status with group as one of the fixed effect. If an effect of pain status or an interaction existed, then rLBP-A and rLBP-R were compared with CTRL separately. The α level was set at 0.05. Statistical analyses were performed in R.^40^ For clarity, here we only present the main effect of pain status or group which is of primary interest, please see supplementary table 1 for step width main effects.

## Results

### Participant characteristics

The rLBP and control groups were matched, hence not different statistically (Table 1). There were significant differences between rLBP-A and rLBP-R for ODI, FABQ-physical activity subscale, pain at rest, and pain during gait (Table 2). Participants with rLBP returned for testing in remission after 47.9 ± 44.2 days, and the last time they recalled having pain was 10 ± 6.9 days ago. Participants with rLBP had pain localized between the 12^th^ rib and gluteal fold (Supplementary figure 2).

**Table 1.** Participant demographics (mean ± standard deviation).

|  | **rLBP (n=20)** | **Control (n=20)** | **P-value** |
| --- | --- | --- | --- |
| **Sex** | 6 M, 14 F | 6 M, 14 F | -- |
| **Age (years)** | 25.3 ± 5.2 | 26.3 ± 3.3 | 0.192 |
| **Height (cm)** | 167.0 ± 7.8 | 165.5 ± 9.9 | 0.534 |
| **Weight (kg)** | 64.2 ± 12.4 | 61.4 ± 12.7 | 0.244 |
| **BMI (kg/m<sup>2</sup>)** | 23.0 ± 4.0 | 22.2 ± 2.8 | 0.132 |
| **Activity (MET)** | 3216 ± 2991 | 2590 ± 1450 | 0.393 |

**Table 2.**
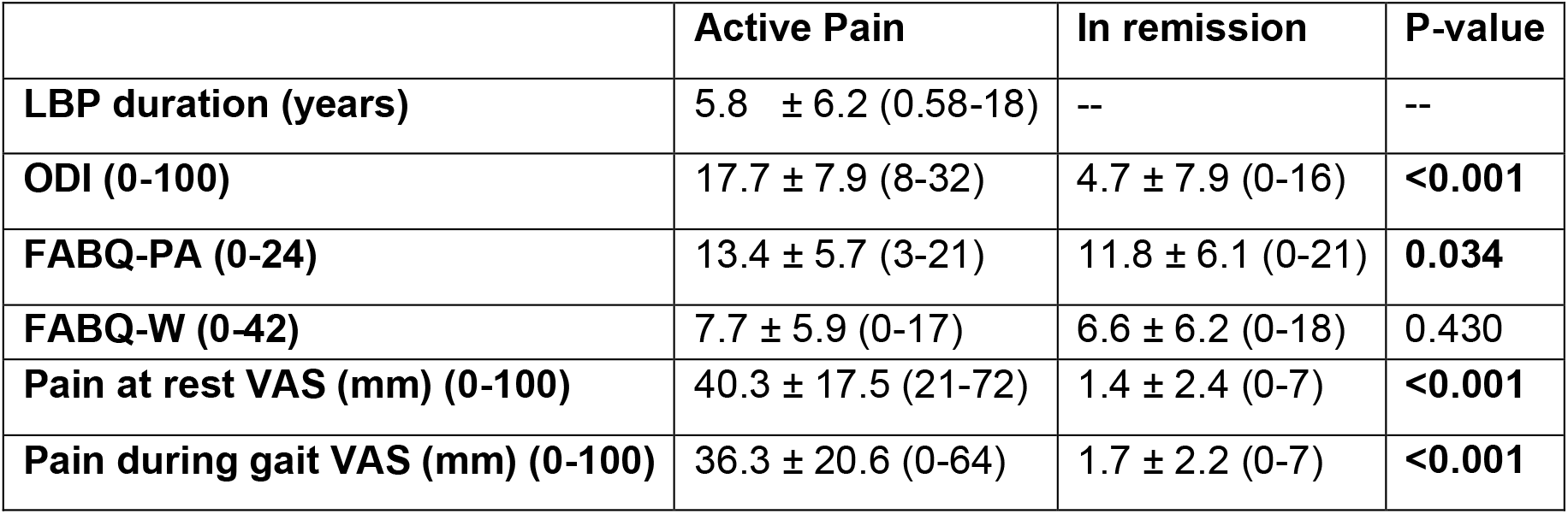
Mean ± standard deviation (range) for low back pain characteristics. ODI: Oswestry Disability Index; FABQ-PA: Fear Avoidance Beliefs Questionnaire – Physical Activity subscale; FABQ-PA: Fear Avoidance Beliefs Questionnaire – Work Subscale; VAS: Visual Analog Scale. Significant p-values are in bold.

|  | **Active Pain** | **In remission** | **P-value** |
| --- | --- | --- | --- |
| **LBP duration (years)** | 5.8 ± 6.2 (0.58-18) | -- | -- |
| **ODI (0-100)** | 17.7 ± 7.9 (8-32) | 4.7 ± 7.9 (0-16) | **<0.001** |
| **FABQ-PA (0-24)** | 13.4 ± 5.7 (3-21) | 11.8 ± 6.1 (0-21) | **0.034** |
| **FABQ-W (0-42)** | 7.7 ± 5.9 (0-17) | 6.6 ± 6.2 (0-18) | 0.430 |
| **Pain at rest VAS (mm) (0-100)** | 40.3 ± 17.5 (21-72) | 1.4 ± 2.4 (0-7) | **<0.001** |
| **Pain during gait VAS (mm) (0-100)** | 36.3 ± 20.6 (0-64) | 1.7 ± 2.2 (0-7) | **<0.001** |

### Task Performance

There were no differences in preferred step width between CTRL, rLBP-A, and rLBP-R (Supplementary figure 3A). Participants varied their step widths based on the visual feedback with only slightly elevated errors in the narrower widths and there were no differences in how well they performed the prescribed widths between CTRL, rLBP-A, and rLBP-R (Supplementary figure 3B).

### Trunk excursion and trunk coordination

There were no significant interactions between step width and pain status or step width and group for any kinematic variables. Regardless of step widths, trunk excursion was significantly higher in rLBP-R compared to both rLBP-A and CTRL (Fig 2). The rLBP-R had reduced in-phase coordination compared to both rLBP-A and CTRL and had greater anti-phase coordination compared to rLBP-A (Fig 3). The rLBP had greater pelvis-only coordination and reduced thorax-only coordination compared to the CTRL group (Fig 3).

**Figure 2.**
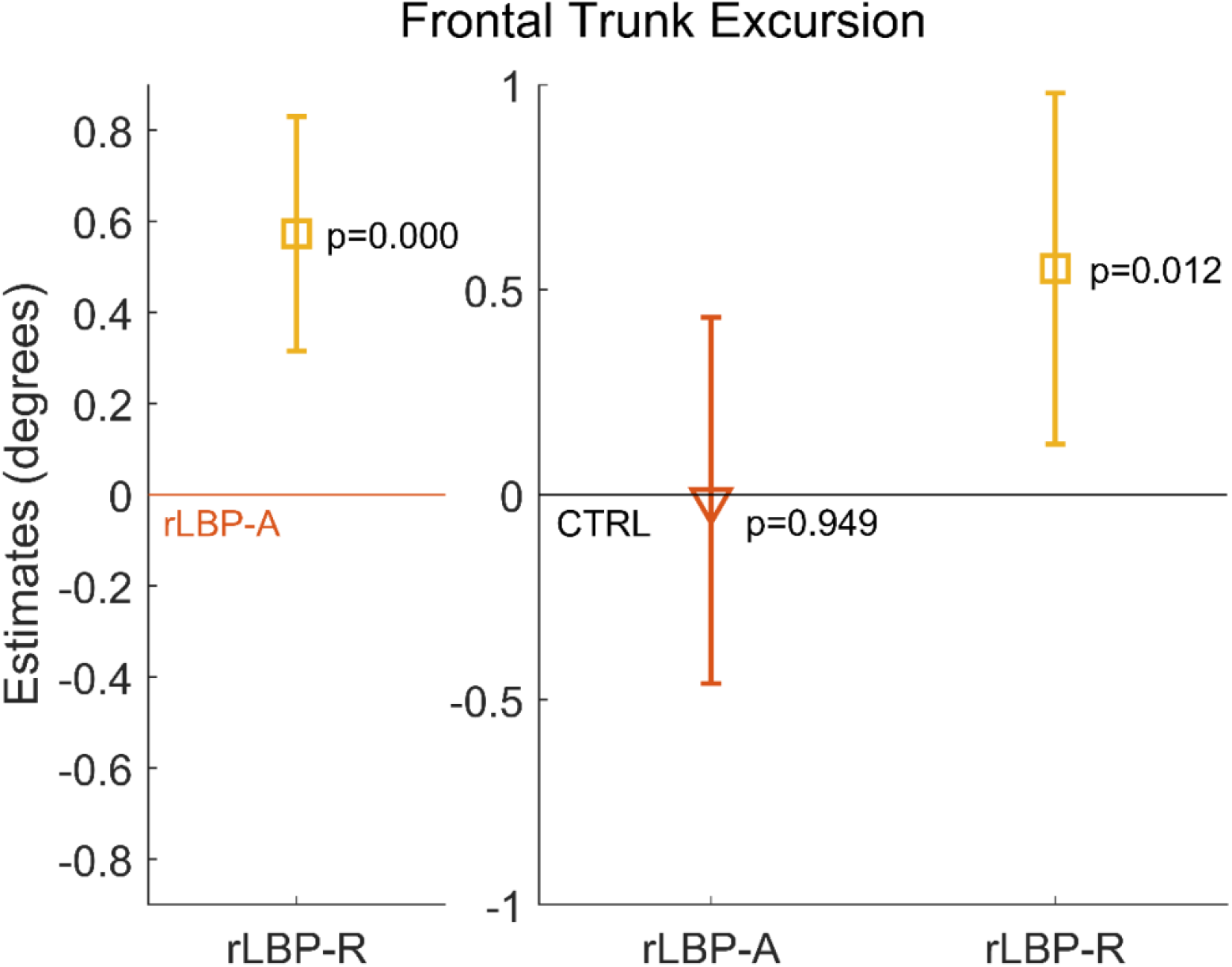
The estimate and 95% confidence interval for the effect of pain (left figure, rLBP-R relative to rLBP-A) and effect of group (right figure, rLBP-A and rLBP-R relative to the CTRL) on frontal plane trunk excursion.

**Figure 3.**
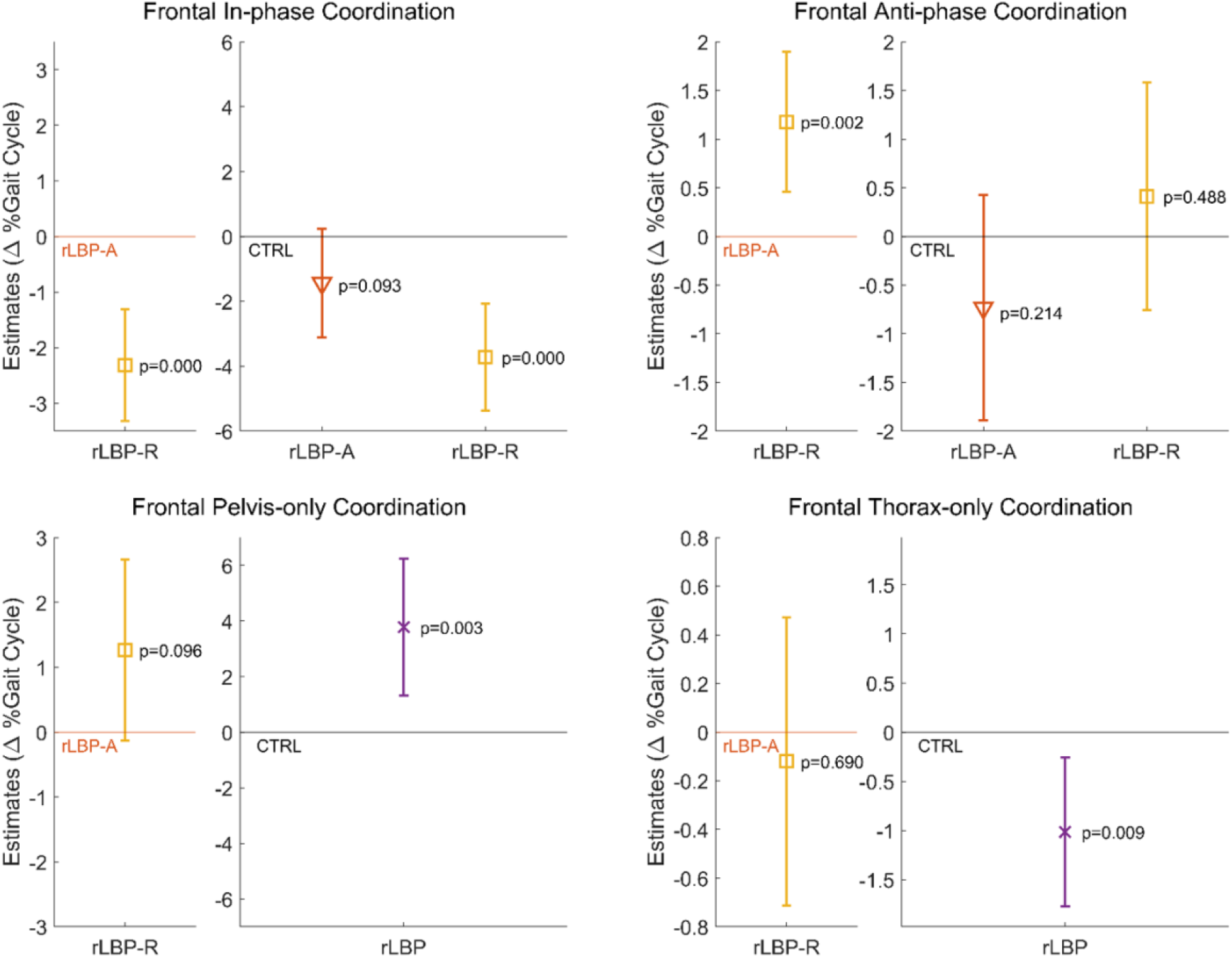
The estimate and 95% confidence interval for the effect of pain (left figure of each plot, rLBP-R relative to rLBP-A) and effect of group (right figure of each plot, rLBP-A and rLBP-R relative to CTRL or rLBP-pooled relative to CTRL) on frontal plane coordination patterns: in-phase, anti-phase, pelvis-only, and thorax-only.

### EMG

There were no significant interactions between step width and pain status or step width and group for any EMG variables. Peak longissimus activation was not different between rLBP-A and rLBP-R, or rLBP and CTRL, but the rLBP had reduced bilateral longissimus co-activation compared to the CTRL (Fig 4).

**Figure 4.**
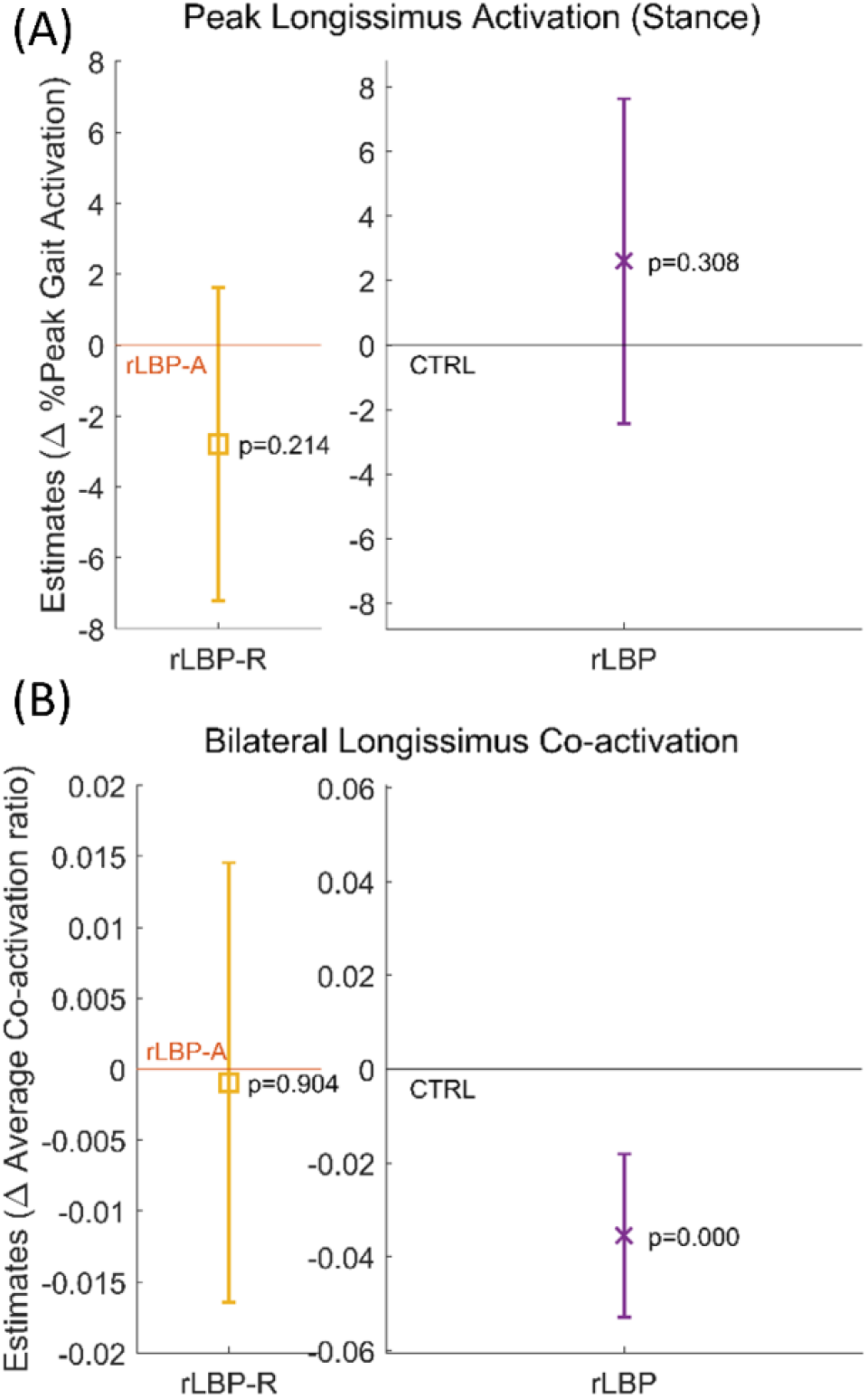
The estimate and 95% confidence interval for the effect of pain (left figure of each plot, rLBP-R relative to rLBP-A) and effect of group (right figure of each plot, rLBP-A and rLBP-R relative to CTRL or rLBP-pooled relative to CTRL) on (A) peak longissimus activation, and (B) bilateral longissimus co-activation.

## Discussion

This study was the first to our knowledge that tested a cohort of young adults with recurrent LBP in and out of a painful episode to determine if alterations in trunk control persist beyond symptom duration. In the subsequent discussion we will use the trunk motor control framework relying on kinematics and muscle activation data. First, we found that participants with recurrent LBP, regardless of pain status, demonstrated a looser instead of a tighter trunk control compared to back-heathy individuals. Second, during symptom remission, participants with recurrent LBP exhibited an even looser control compared to when they were in pain. Inconsistent with our expectation of altered adaptability, participants with recurrent LBP did not differ from back-healthy individuals in their ability to modify trunk control in response to different step widths.

Focusing first solely on the effect of pain status on trunk control in participants with recurrent LBP, active pain resulted in trunk kinematics consistent with tighter control compared to when participants were in remission. Pain status, however, did not affect longissimus muscle activation. This tightened behavior could be an attempt to protect the region from unexpected segmental motion, and was consistent with previous literature in clinical and experimental LBP.^19,20,41–44^ Based on our findings, the alterations in trunk kinematics in and out of pain was not due to changes in peak longissimus activation and bilateral longissimus co-activation; alternatively, they could be due to changes in other trunk muscle activities and deep-to-superficial muscle coordination.^11,44^

It is likely that varying task demands reveal different movement strategies in people with LBP.^17,18^ The majority of previous experimental studies suggest individuals with LBP present with tighter trunk control than back-healthy persons. These studies utilize fast, discrete perturbations such as a sudden load^45^, unpredictable weight release,^19,46^ or posteroanterior thrust on the spine,^41^ and the results may reflect neuromuscular reflexive responses. Studies using fast, large perturbations, however, may have limited ecological validity since people report that their LBP recurrence typically occurs during submaximal physical activities.^47^ On the other hand, studies utilizing self-initiated tasks reflect existing motor programs and often demonstrate a looser trunk control in persons with clinical LBP compared to controls. Increased lumbar excursion during the early phase of picking up an object^27,48^, increased angular displacement at specific lumbar segments during trunk flexion^49^, and decreased trunk coupling during a continuous spring-compression task^50^ have all been demonstrated in individuals not in an acute flare-up, but in remission of chronic or recurrent LBP. While perturbation-type studies may be tapping into neuromuscular reflex responses, studies with self-initiated tasks observe the voluntary movement patterns that has been established and utilized by the participants. It have been hypothesized that individuals with LBP use increased co-activation to compensate for delayed feedback and aberrant sensory input when facing perturbations.^51^ On the other hand, segmental hypermobility^52^ and decreased muscle force due to fat infiltration and/or atrophy^53^ may contribute to the looser trunk control associated with LBP.

We showed that participants with recurrent LBP exhibited looser trunk kinematics and lower muscle activation during walking with various step widths compared to the back-healthy controls, regardless of pain status. This was more pronounced during symptom remission, and even when they tightened their control during a painful episode, they demonstrated a looser strategy than the control group. Previous studies on walking gait had inconsistent results on trunk kinematics and coordination, showing either tighter^20,24,42,43,54–56^, looser^21,56^, or no difference^43,56,57^ in trunk control in participants with a history of LBP compared to back-healthy individuals. These studies include individuals who were symptomatic^20,24,42,43,54–57^ and asymptomatic^20,21,24^ at the time of testing, but there was no clear indication of pain status corresponding to tighter or looser control. Walking is a self-initiated, continuous, and non-provocative task that is well practiced, therefore suitable for examining habitual movement patterns in persons with LBP. Our results were consistent with other studies using self-initiated tasks and suggest that individuals with recurrent LBP exhibit looser trunk control than back-healthy persons, and these changes may exist prior to the LBP episode we tested. We must be mindful that studies used various methods of computing trunk coordination, reported results in different planes of motion, and adopted diverse definitions of the LBP cohort. Before we can attribute a loose or tight behavior to a patient, much more patient-specific assessment is needed.

The further altered trunk control during symptom remission may suggest that movement patterns or anatomical factors (e.g. pelvis morphology or ligament laxity) existing prior to the tested painful episode underlie the motor behavior found in individuals with recurrent LBP. This interpretation is distinct from a previously popular perspective that altered motor control was an unresolved behavior from the painful periods, in which case we should see less or similar aberrant movement during symptom remission than in a painful episode. The longitudinal design of this study allowed us to uncover the nature of altered motor control in the context of recurrent pain that may have otherwise been missed.

Our study has some limitations. To maximize feasibility, we tested participants first in pain and then out of pain. Therefore, participants may have performed differently out of pain simply because of previous exposure to the task. We performed test-retest comparisons and did not find evidence indicating previous exposure having an effect (Supplementary figure 4). Additionally, despite good to excellent test-re-test reliability in the control participants^32^, reliability in individuals with recurrent LBP should be examined as it may not be the same as the back-healthy controls.

In conclusion, young individuals with mildly disabling recurrent LBP exhibited looser trunk kinematics and lesser muscle activation strategies compared to back-healthy controls, regardless of pain status. These strategies included decreased bilateral longissimus co-activation and greater pelvis-only coordination in the frontal plane during gait. This looser trunk control strategy was further exaggerated when they were in symptom remission, demonstrated by an increased trunk excursion, decreased in-phase and increased anti-phase coordination. Future studies should explore if the trunk control alterations seen in active pain or during remission can predict future symptoms through a longitudinal design to uncover opportunities for designing targeted, effective interventions.

## Data Availability

Data may be available, please contact the corresponding author.

## Acknowledgements

This research was supported by the International Society of Biomechanics Matching Dissertation Grant, with matching contribution from the USC Division of Biokinesiology and Physical Therapy.

---

**Supplementary table 1.**
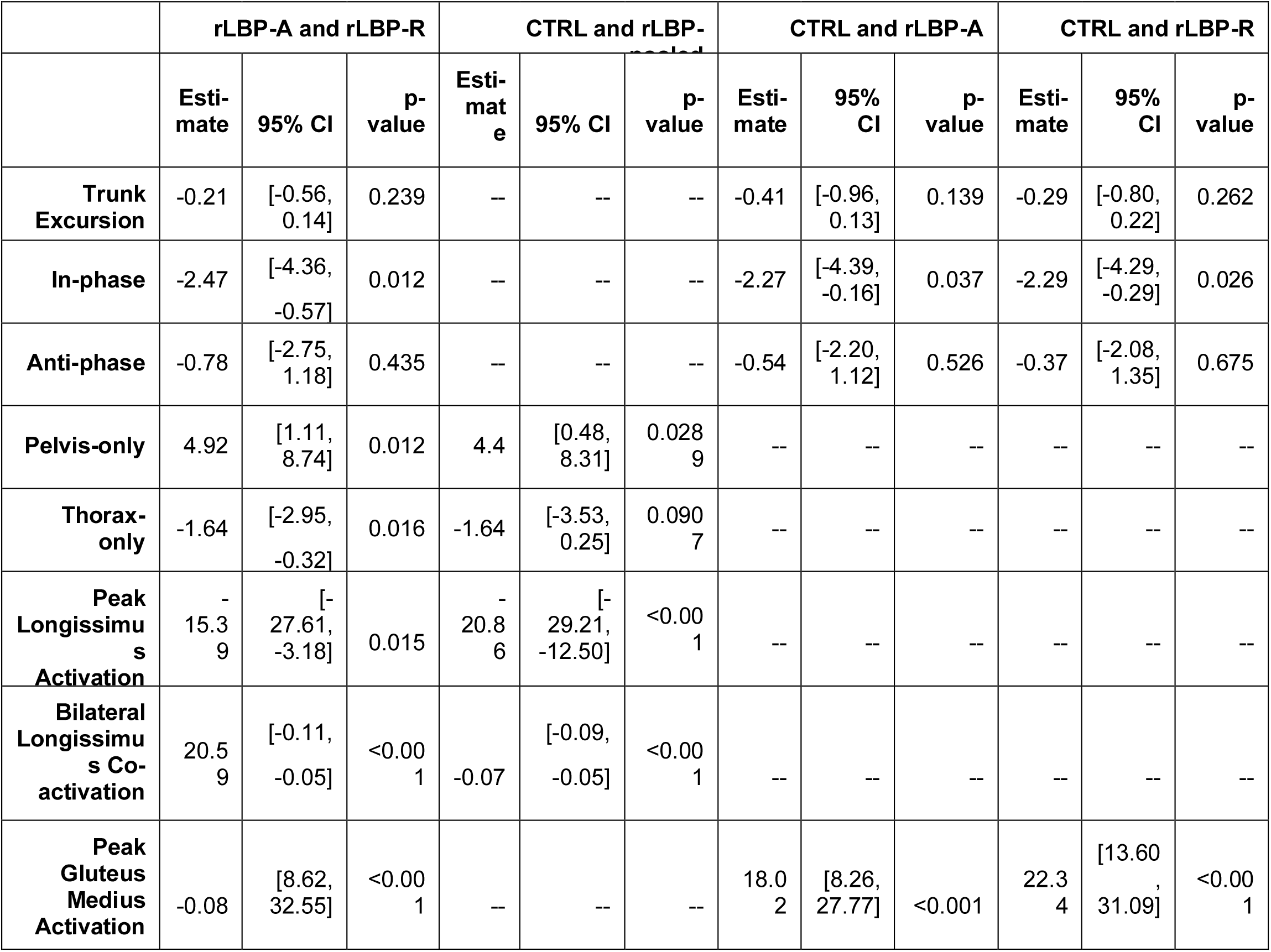
Step width effects statistical results from general mixed-effects models. CTRL: Back-healthy controls; rLBP-A: recurrent low back pain group in active pain; rLBP-R: recurrent low back pain group in remission; rLBP-pooled: pooled data across two testings for recurrent low back pain group (only when there was no pain effect between

## Supplementary Figures

**Supplementary figure 1.**
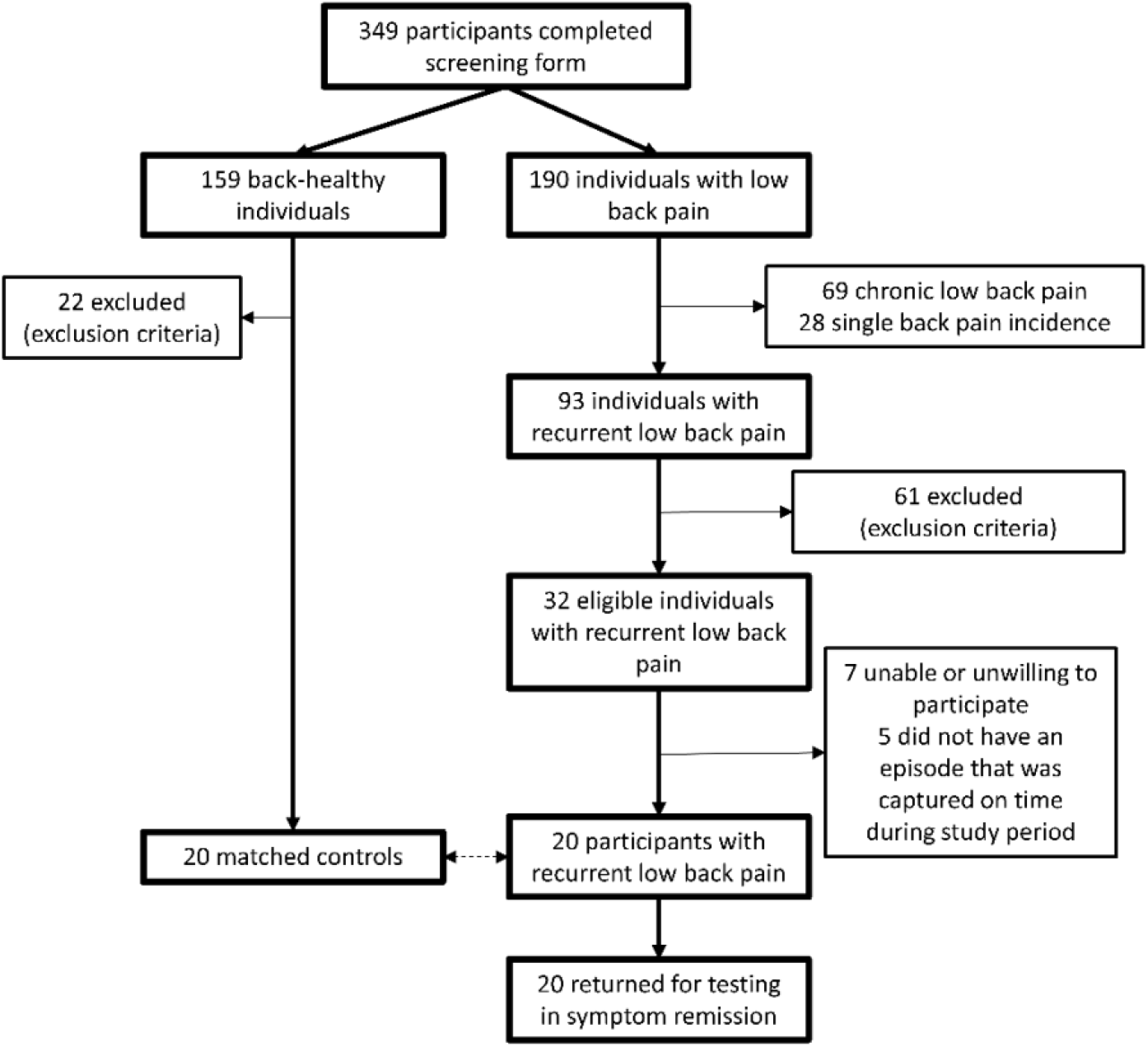
Participant consort diagram.

**Supplementary figure 2.**
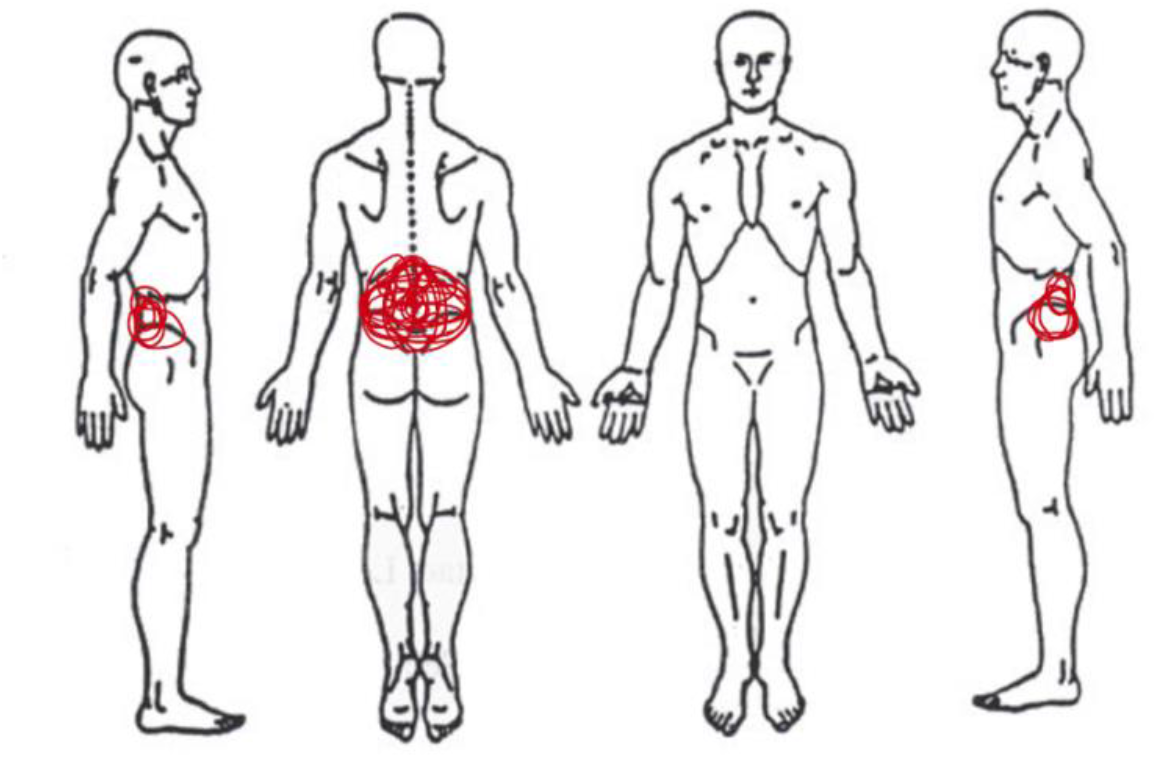
Self-reported body pain diagram composite indicating rLBP participants’ primary pain location during the active pain testing session.

**Supplementary figure 3.**
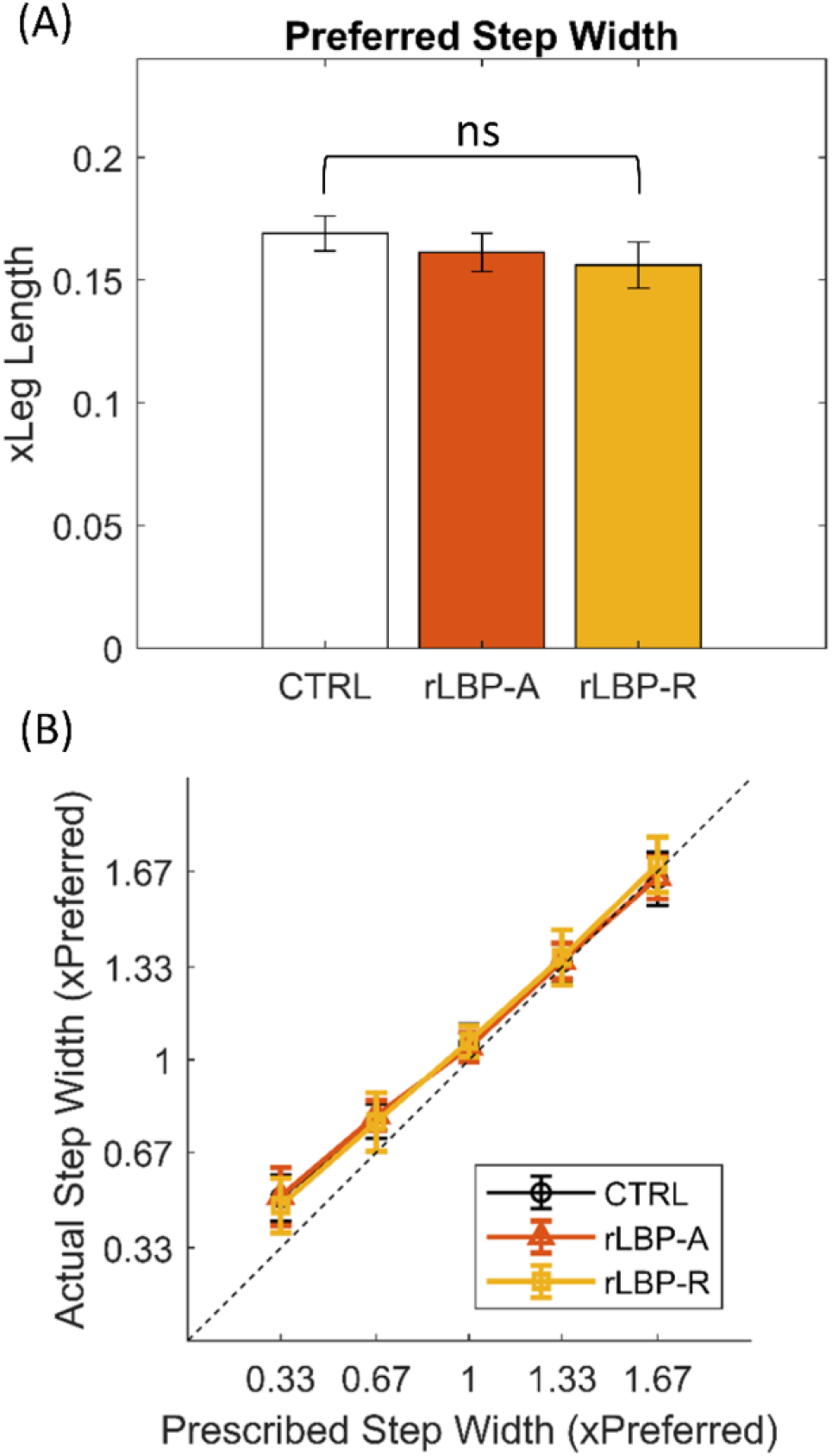
Step width task performance. Both plots showing data for the back-healthy controls (CTRL), individuals with recurrent low back pain while in active pain (rLBP-A), and the same individuals during symptom remission (rLBP-R). (A) Mean and standard error of mean of preferred step width (B) Square plot presenting the mean and standard deviations of actual step width performance relative to the prescribed step widths. Note that the 1 x preferred step width was also prescribed with visual feedback. The diagonal reference line indicates a one-to-one fit of the performance with the targets.

**Supplementary figure 4.**
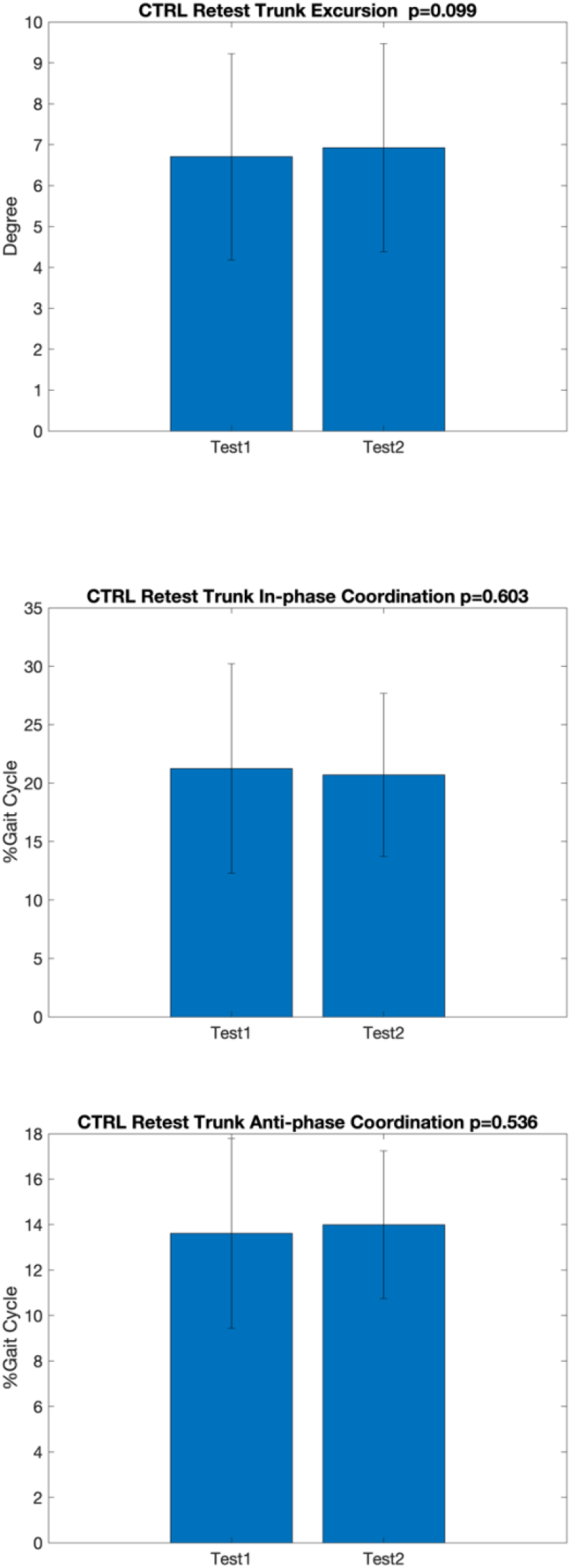
We compared test-retest performance using paired t-tests on 6 control participants re-tested a week apart for any variable that was significantly different between rLBP-A and rLBP-R to examine the possibility that the difference was merely due to previous exposure to the task. No differences between test-retest for trunk excursion, and trunk in-phase and anti-phase coordination was found, indicating no evidence of an effect of previous exposure to the experimental task.

## Notes

### Competing Interest Statement

The authors have declared no competing interest.

### Author Declarations

University of Southern California Health Science Campus IRB #HS-16-00980.

